# Short-Term Modulation of Epileptic Network with Low-Frequency Thalamic Stimulation

**DOI:** 10.1101/2025.10.27.25338922

**Authors:** Sotaro Kanai, Atsuro Daida, Saarang Panchavati, Roshan Srinivasan, Tonmoy Monsoor, Joe X Qiao, Noriko Salamon, Raman Sankar, Aria Fallah, Jerome Engel, Richard J Staba, Vwani Roychowdhury, William Speier, Hiroki Nariai

## Abstract

Thalamic neuromodulation is an emerging therapeutic strategy for patients in whom resection of the epileptogenic zone is not feasible. Yet the mechanisms by which thalamic stimulation modulates epileptogenic network dynamics remain unclear. We designed this study to test whether brief thalamic stimulation transiently modulates cortico-cortical activity and coupling within patient-specific epileptic networks.

We studied 10 patients undergoing stereo-EEG with sampling of the anterior or centromedian thalamic nuclei. During wakefulness, we delivered low-frequency single pulse stimulations at 1 Hz and compared post-stimulation epochs (15–900 ms) with a pre-stimulation baseline. We quantified band-limited spectral power and Granger causality between bipolar cortical contacts, focusing on slow (3.0–13.9 Hz) and fast (14.0–79.9 Hz) activity. Contacts were grouped as seizure onset zone, early-propagating regions, or other regions, emphasizing the hemisphere ipsilateral to the stimulated nucleus.

Thalamic stimulation produced a rapid, distributed modulation of cortical dynamics. Across patients, power in early-propagating regions decreased consistently in both slow and fast bands, whereas seizure onset zone and other regions showed smaller and less consistent changes, indicating reduced recruitment of cortical regions that often participate in seizure propagation. Directed interactions measured by Granger causality increased modestly at the network level, with the most consistent change between the seizure onset zone and other regions, and a significant increase in influence directed toward the seizure onset zone. These effects were observed with both nuclei despite heterogeneous implant strategies, suggesting a conserved, thalamus-initiated rebalancing of cortico-cortical coupling. Notably, when the anterior nucleus was stimulated in patients with limbic seizure onset zone, the power decrease in early-propagating regions and the increase in directed influence between the seizure onset zone and other regions were prominent, consistent with anterior thalamic projections to the limbic network.

Taken together, these findings point to a rapid, nucleus-specific modulation that engages thalamic nodes while recruiting broader cortico-cortical influence toward the seizure onset zone. The sub-second markers—characterized by reduced power in early-propagating regions and selective increases in directed influence between seizure onset zone and other regions— support a model in which thalamic input modulates cortico-cortical coupling to limit propagation of epileptic activity from seizure onset zone to early-propagating regions. By characterizing network effects on the timescale of therapeutic stimulations, this work identifies mechanistic markers for network-informed neuromodulation, where transient EEG responses can guide nucleus selection and parameter titration. Prospective studies that link these markers to long-term seizure outcomes and stratify by thalamocortical anatomy are warranted.

## Introduction

Approximately 25–40 % of epilepsy patients develop drug-resistant epilepsy (DRE).^1,2^ When the presumed epileptogenic zone (EZ) is clearly defined and localized to non-eloquent areas, resection or disruption can yield seizure freedom. In contrast, complete removal of the EZ is often precluded when it is multifocal or involves eloquent cortex, limiting available treatment options.^1,2^

To address this therapeutic gap, thalamic neuromodulation has emerged as a key therapeutic option. Deep brain stimulation (DBS) of the anterior nucleus (AN) of the thalamus reduces seizures with durable long-term benefit in focal DRE.^3,4^ In patients with multifocal or generalized phenotypes, DBS and brain-responsive neurostimulation (RNS) targeting the centromedian (CM) nucleus in patients with multifocal or generalized phenotypes likewise demonstrates progressive, sustained seizure reduction in prospective follow-up.^5–9^ Thalamic RNS has also shown benefit in generalized epilepsies, although rigorous selection criteria remain unsettled.^10–12^ Although thalamic DBS and RNS show increasing clinical efficacy in DRE, human causal evidence showing how thalamic stimulation modulates the epileptogenic network over short and long timescales remains scarce. Few studies have combined thalamic stimulation with intracranial recordings, leaving the mechanisms of network modulation and therapeutic benefit unresolved.

A mechanistic explanation may be derived from thalamocortical network physiology. The thalamus actively regulates functional interactions within and across cortical systems, and higher-order thalamic nuclei can rapidly reconfigure large-scale cortical coupling.^13^ In parallel, epilepsy is widely conceptualized as a disorder of networks, with seizure initiation, propagation, and termination mediated by dynamic, directed interactions across vulnerable hubs.^14,15^ Human intracranial and multi-nuclear recordings show early thalamic recruitment and thalamocortical synchrony that track seizure propagation and generalization, with AN and CM coupling to limbic and fronto-parietal circuits linked to ictal dynamics.^16–21^ Together, these perspectives suggest that therapeutic thalamic stimulation may act by modulating network excitability and coupling along thalamocortical loops.

Direct intracranial recordings have begun to clarify these dynamics in humans. Using stereo-EEG (SEEG) with thalamic sampling, studies show that corticothalamic connectivity increases around ictal onset with a frequency-specific bias towards slower bands, while thalamic outflow exhibits distinct spectral structure.^22,23^ Multisite thalamic SEEG demonstrated early and nucleus-specific thalamic involvement and mapped how stimulation-evoked activity follows intrinsic thalamocortical networks.^17,18^ In pediatric cohorts, thalamic SEEG has demonstrated thalamocortical network activation during epileptic spasms.^24^ Together, these findings motivate systematic investigations using controlled thalamic stimulation paired with intracranial network analysis to delineate how stimulation engages and modulates the epileptic network.

Single-pulse electrical stimulation (SPES) during intracranial monitoring offers a direct way to test effective connectivity and cortical excitability, yielding cortico-cortical evoked potentials (CCEPs) that are now used to map pathways and assess excitability under established safety parameters.^25,26^ While SPES-based mapping of cortico-cortical networks is mature, studies targeting thalamic nuclei and quantifying downstream network effects remain limited but are rapidly expanding: stimulation of human AN evokes cortical responses that align with intrinsic functional connectivity, pointing to propagation along pre-existing thalamocortical networks^18^; multi-nucleus perturbation–recording datasets provide an emerging atlas of causal thalamocortical connections^27^; and prolonged thalamic stimulation during SEEG has been shown to suppress interictal spikes and reduce evoked-potential amplitudes in connected cortices, consistent with network-level reductions in excitability.^28^

Guided by these insights, we hypothesized that brief thalamic stimulation would rapidly modulate the epileptogenic network, attenuating propagation of activities from SOZ to the surrounding cortex. In patients undergoing SEEG with thalamic coverage, we therefore used SPES as a direct way to test network effects and quantified post-stimulation changes in band-limited power and directed interactions via Granger causality (GC) within and around the SOZ, with concordance assessed on simultaneous scalp EEG. We show frequency-specific reductions of cortical power along with suppression of propagation-related features and reorganization of directed cortical interactions, supporting a mechanistic link between thalamic neuromodulation and seizure control.

## Materials and methods

### Patient cohort

We screened all patients diagnosed with drug-resistant epilepsy (DRE) before age 18 who underwent video-SEEG monitoring for epilepsy surgery at UCLA Mattel Children’s Hospital between January 2023 and May 2025. Among these, 11 had at least one thalamic SEEG electrode and underwent SPES during monitoring. One patient was excluded due to pervasive, non-removable recording artifacts, yielding a final analytic cohort of 10 (5 males, 5 females). Median age of epilepsy onset was 3.5 years, and median age of SEEG implantation was 16 years. All patients had focal epilepsy except one with Lennox–Gastaut syndrome and concurrent focal seizures. Etiologies were structural (n=4), genetic (n=2), immune (n=2), and unknown (n=2). The study protocol was approved by the UCLA institutional review board. Written informed consent was obtained from all patients and/or their guardians. All clinical evaluations were part of standard care, and EEG data were de-identified before analysis. All included patients had SEEG implantation and complete imaging (pre-implant T1-weighted MRI and post-implant CT) as well as SEEG recordings spanning the SPES sessions. Patient demographics, clinical characteristics, and electrode information are summarized in **Table 1**.

**Table 1.** Cohort characteristics.

| Pt No. | Age at SEEG (years) | Age at epilepsy onset (years) | Etiology | Epilepsy type | Seizure type | Prior surgery | ASM at stimulation | Weaned ASM at stimulation | No. of SEEG contacts placed | Bilateral coverage | Thalamic nuclei | SOZ side | SOZ lobe | Number of SOZ channels | Number of EARLY channels | Covered lobe | No. of analyzed thalamic stimulation channels | No. of excluded thalamic stimulation channels | No. of analyzed cortical channels | No. of excluded ipsilateral cortical channels | No. of contralateral channels | Scalp EEG | Surgery after SEEG | Thalamic RNS |
| --- | --- | --- | --- | --- | --- | --- | --- | --- | --- | --- | --- | --- | --- | --- | --- | --- | --- | --- | --- | --- | --- | --- | --- | --- |
| 1 | 0–4 | 0–4 | Encephalomalacia | Focal | FIAS | None | CLB, LTG, PER | N | 94 | Y | L: CM, PUL | L | O | 3 | 3 | F,P,O,T,LIM | 2 | 0 | 28 | 3 | 47 | Y | RNS | Y |
| 2 | 5–9 | 0–4 | TSC | Focal | FIAS, ES | None | LEV, LTG, VGB | N | 98 | Y | L: CM<br>R: CM | R | LIM | 2 | 32 | F,P,T,LIM | 4 | 2 | 34 | 4 | 41 | Y | Right multiple tuber resection | N |
| 3 | 10–14 | 5–9 | TSC | Focal | FIAS | None | CNB, LTG, ZNS | N | 56 | Y | L: AN, CM | L | F | 3 | 0 | F,P,T,LIM | 2 | 2 | 22 | 5 | 15 | Y | Left orbitofrontal tuber resection | N |
| 4 | 15–19 | 0–4 | Unknown | Focal | FAS | None | CLB, LCM, LEV | N | 114 | Y | L: AN, CM | Bil* | F, LIM | 6 | 8 | F,T,LIM | 4 | 0 | 42 | 11 | 41 | N | RNS | Y |
| 5 | 15–19 | 5–9 | Unknown | Focal | FBTCS | None | BRV, CLB, CNB | All off | 98 | Y | R: AN, CM, PUL | R | F, LIM | 12 | 7 | F,T,LIM | 4 | 0 | 46 | 9 | 21 | Y | RNS | N |
| 6 | 15–19 | 0–4 | Band heterotopia | LGS | FMS, ES, TS | VNS | CLB, CNB, RUF | N | 48 | Y | L: AN, CM, PUL<br>R: AN, CM, PUL | Bil | O | 9 | 13 | O | 3 | 4 | 22 | 0 | 0 | Y | RNS | Y |
| 7 | 15–19 | 10–14 | Autoimmune encephalitis | Focal | FMS | VNS | CLB, CNB, ZNS | ZNS off, CNB half | 80 | Y | L: AN, CM<br>R: AN, CM | R | F, LIM | 6 | 4 | F,T,LIM | 4 | 4 | 35 | 8 | 20 | Y | RNS | N |
| 8 | 20–24 | 0–4 | FCD | Focal | FIAS, FBTCS | None | CLB, LTG | CLB half, LTG half | 94 | Y | R: AN, CM | R | F, T | 7 | 2 | F,T,LIM | 3 | 1 | 35 | 8 | 34 | Y | None | N |
| 9 | 20–24 | 5–9 | FCD | Focal | FIAS | RNS | BRV, CNB, ESL, PER | N | 90 | Y | L: AN<br>R: CM | L | F,T,LIM | 7 | 14 | F,T,LIM | 3 | 3 | 41 | 24 | 9 | N | RNS | N |
| 10 | 20–24 | 0–4 | Unknown | Focal | FMS | Right ATL | BRV, CNB, LCM | N | 93 | N | R: AN, CM, PUL | R | T | 3 | 13 | F,P,O,T,LIM | 4 | 0 | 60 | 12 | 0 | Y | RNS | Y |

\* The right SOZ was not analyzed due to the absence of ipsilateral thalamic coverage in this patient. Due to ethical considerations, ages are given in ranges.
Abbreviations: SEEG = stereo-EEG; ASM = anti-seizure medication; SOZ = seizure onset zone; EARLY = early-propagating region; DBS = deep brain stimulation; RNS = responsive neurostimulation; TSC = tuberous sclerosis complex; FCD = focal cortical dysplasia; LGS = Lennox-Gastaut syndrome; FIAS = focal impaired awareness seizure; ES = epileptic spasms; FAS = focal aware seizure; FBTCS = focal to bilateral tonic clonic seizure; FMS = focal motor seizure; TS = tonic seizure; VNS = vagus nerve stimulation; ATL = anterior temporal lobectomy; BRV = brivaracetam; CLB = clobazam; CNB = cenobamate; ESL = eslicarbazepine; LCM = lacosamide; LTG = lamotrigine; PER = perampanel; RUF = rufinamide; ZNS = zonisamide; AN = anterior nucleus; CM = centromedian nucleus; PUL = pulvinar nucleus; F = frontal; O = occipital; T = temporal; LIM = limbic (hippocampus + amygdala + cingulate gyrus + insula + orbitofrontal cortex).

### Patient evaluation

All patients with DRE underwent a standardized presurgical work-up comprising scalp video-EEG monitoring, high-resolution (3.0T) brain MRI, and ^18F-fluorodeoxyglucose-PET with MRI-PET co-registration. SEEG placement was planned through a multidisciplinary epilepsy surgery conference that integrated seizure semiology, neurological examination, structural and functional neuroimaging, neuropsychological evaluation, and scalp EEG, with emphasis on seizure onset. Thalamic SEEG coverage was selected for patients with multifocal or unclear SOZs, those exhibiting both focal and generalized seizures, and/or when candidacy for thalamic neuromodulation—DBS or RNS— was under consideration at the conference.^8,22–24^ Seven of the ten patients subsequently underwent RNS implantation, including four who received thalamic RNS leads.

### iEEG placement and patient-specific brain modelling

SEEG depth macroelectrodes were surgically implanted with 2.5-mm contact spacing for thalamic trajectories and 5-mm spacing for other targets. Trajectory planning was performed in Brainlab Elements™ (Brainlab AG, Munich, Germany) using preoperative T1-weighted and gadolinium-enhanced MRI to guide target and entry selection. Through 2023, implantations were executed using Leksell frame coordinates generated in Brainlab Elements; thereafter, procedures were performed with the ROSA^®^ robotic surgical assistant (Zimmer Biomet, Montpellier, France). For thalamic targeting, an experienced radiologist (N.S.) delineated nuclei of interest, and trajectories were planned in Brainlab Elements. Unilateral or bilateral sampling of the AN, CM, and pulvinar was considered, guided by known anatomical connectivity to the suspected SOZ and to identify potential neuromodulation targets.

**Fig. 1** illustrates the analytical workflow. For each patient, a preoperative high-resolution 3D T1-weighted MRI was acquired, and patient-specific 3D brain models were generated using FreeSurfer.^23,24^ Intraoperative or postoperative CT was obtained to rule out intracranial hemorrhage and to confirm electrode positions. Thin-cut CT volumes were co-registered to the 3D brain models using Brainstorm software, enabling precise localization of each electrode contact in native space. Cortical SEEG contact labels were assigned based on the Desikan-Killiany-Tourville (DKT) atlas. Thalamic contact locations were verified using FreeSurfer-based automated segmentation. Contacts whose centers lay within a given nucleus, or within 2 mm of its boundary, were considered analytic candidates for AN and CM. Pulvinar contacts were excluded from analysis because the sample size was insufficient to support group-level inference.

**Fig. 1.**
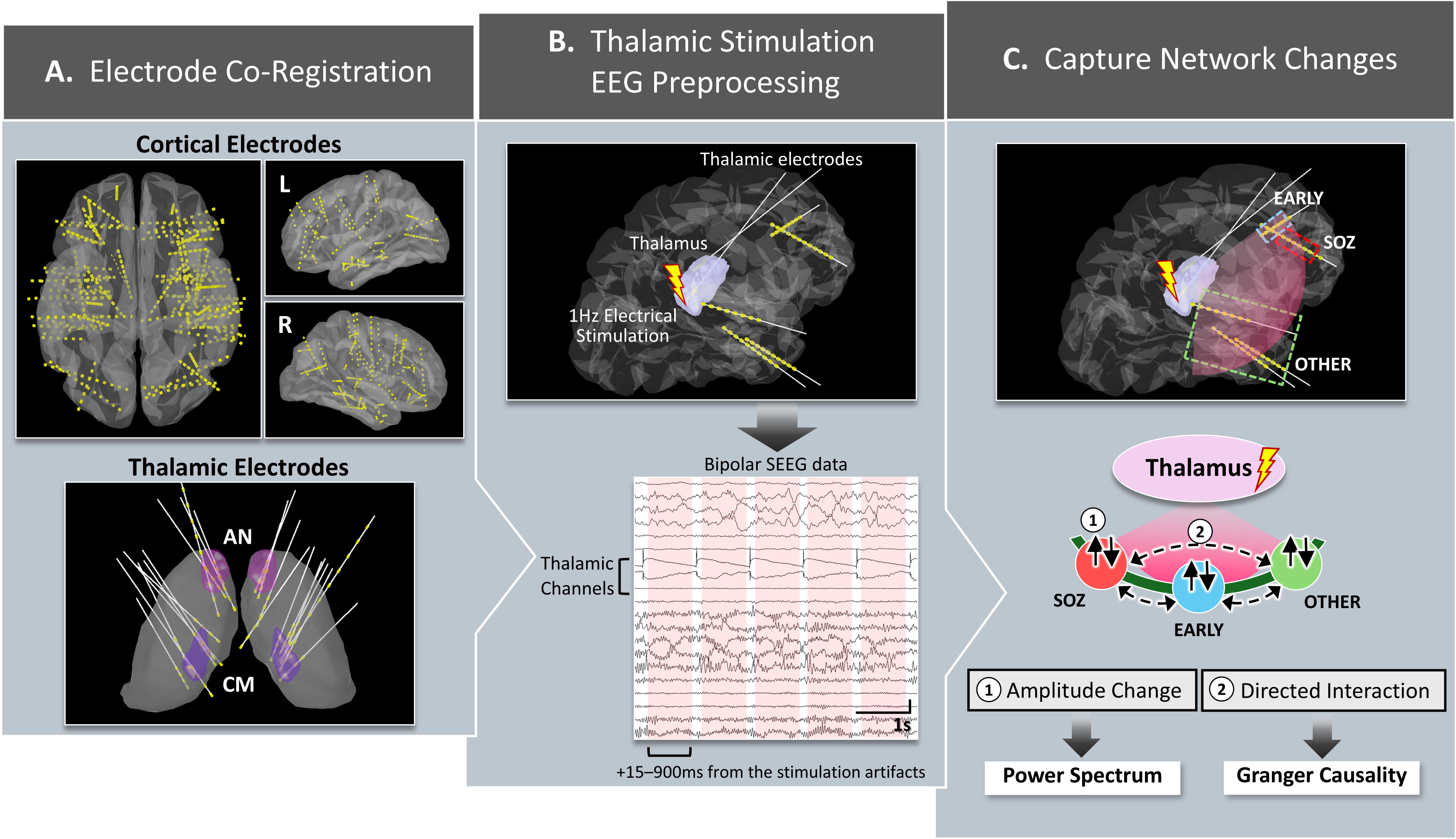
Study design and analysis overview. (A) Electrode co-registration and channel taxonomy. Pre-implant T1-weighted MRI and post-implant CT were co-registered to localize SEEG contacts, including thalamic leads targeting AN and CM. Cortical bipolar channels were classified as SOZ, early-propagation (EARLY), or other cortex (OTHER); analyses were restricted to contacts in the hemisphere ipsilateral to the stimulated thalamus. (B) Thalamic SPES. Bipolar thalamic pairs were stimulated at 1 Hz with single 0.2-ms pulses, delivered sequentially across predefined contact pairs. The stimulation artifact defined time zero; post-stimulus analysis windows spanned +15–900 ms per pulse. (C) Signal processing and network metrics. SEEG was analyzed in a bipolar montage; scalp EEG (when available) used an average reference. Band-limited power was computed per epoch and summarized primarily in slow (3.0–12.9 Hz), fast (13.0–79.9 Hz), and high-frequency activities (80.0– 299.9 Hz) bands. Directed interactions were estimated with time-domain Granger causality between bipolar SEEG channels; edge-wise changes from baseline were aggregated within and between SOZ, EARLY, and OTHER groups to obtain group-level summaries. SEEG = stereo-EEG; AN = anterior nucleus; CM = centromedian nucleus; SOZ = seizure onset zone.

### Thalamic SPES protocol

On the day after SEEG implantation, we performed cortical SPES to provoke habitual seizure events and refine localization of the epileptogenic zone, while minimizing the risk of prolonged or generalized seizures. The thalamic nuclei were also stimulated in anticipation of potential future neuromodulation therapies, particularly to assess possible stimulation-related side effects. Stimulation was delivered in a bipolar configuration between predefined contact pairs. Each trial consisted of 40 repetitions of a single 0.2-ms pulse at 1 Hz (inter-stimulus interval of 1 s), delivered sequentially to bipolar electrode pairs— excluding any contacts located outside the brain parenchyma. To account for the shorter intercontact spacing in thalamic leads (2.5 mm) and for safety, thalamic pairs were tested at 1 and 2 mA, whereas non-thalamic pairs were tested at 2 and 3 mA. Throughout stimulation, the EEG was monitored for afterdischarges and clinical signs; no thalamic stimulation produced afterdischarges or provoked seizures.

We identified 47 candidate thalamic bipolar stimulation pairs; 14 were excluded because contacts lay outside AN or CM, or because no SOZ channels were present in the ipsilateral hemisphere, leaving 33 stimulated pairs for analysis (2–4 pairs for each patient).

### EEG signal processing

SEEG and simultaneous scalp EEG data were recorded using Nihon Kohden Systems (Irvine, California, USA) at a sampling frequency of 2,000 Hz. All baseline and thalamic-stimulation recordings were obtained during wakefulness. To mitigate volume conduction, SEEG signals were transformed into a bipolar montage, whereas scalp EEG was referenced to the average reference; 60-, 120-, 180-, 240-, and 300-Hz power-line noise was removed with notch filtering. Frequency content was analyzed a priori in three bands—slow (3.0–13.9 Hz), fast (14.0–79.9 Hz), and high-frequency activities (HFA; 80.0–299.9 Hz)— with band specific results reported in the corresponding metric sections.

Bipolar SEEG channels were classified as SOZ channels (SOZ), defined by rhythmic ictal-onset activity agreed upon by the clinical team; early-propagating channels (EARLY), showing spread within 10 s of onset; and other channels (OTHER). Cortical channels were excluded if (1) either contact was outside the brain parenchyma, (2) significant artifacts were present, or (3) the bipolar midpoint lay within 5 mm (Euclidean distance) of the stimulated thalamic contact’s midpoint. To approximate activity from relatively normal cortex, OTHER channels were additionally excluded if (a) both contacts lay in white matter, (b) either contact overlapped lesional cortex (e.g., cortical tuber or heterotopia), or (c) interictal spikes (≥1 per 10 s) were observed. In total, 370 cortical bipolar channels (22–60 per patient) were analyzed, while 307 channels were excluded. Cortical channels were further assigned, based on the DKT atlas, to frontal (F), parietal (P), occipital (O), temporal (T), or limbic (LIM— hippocampus, amygdala, cingulate gyrus, insula, and orbitofrontal cortex) groups. Only channels within the hemisphere ipsilateral to the stimulated thalamus were analyzed; when no ipsilateral Early channels were present for a given thalamic stimulation channel, analyses were limited to SOZ and OTHER. Detailed electrode information is summarized in **Supplementary Table 1**. Given the lower spatial resolution of scalp EEG, scalp channels were grouped relative to the stimulated thalamus as ipsilateral, contralateral, or midline (Fz, Cz, Pz).

The timing of thalamic stimulation was identified from the stimulation artifact. To avoid artifact contamination, post-stimulation analysis windows spanned +15 to +900 ms relative to each artifact (885 ms per epoch). Because of stimulation artifacts, all thalamic bipolar pairs were excluded from power and network analyses. For the baseline, a 35.4-s segment ending 10 s before the first stimulus of a series was partitioned into forty 0.885-s epochs, matching the number and length of post-stimulation epochs. An experienced EEG reader (S.K.) visually reviewed all channels and epochs, and those with marked artifacts were excluded. For each patient, metrics were computed per epoch and then averaged across epochs within condition (baseline vs. post-stimulation); results were subsequently aggregated by stimulated thalamic nucleus and by band (slow/fast/HFA as applicable).

### EEG power spectral analysis

Band-limited power was estimated on the pre-defined 885-ms epochs using Welch’s method, with the fast Fourier transform length matched to the epoch duration and mean averaging across segments. For each channel and condition (baseline vs post-stimulation), power was computed separately for the slow and fast bands and averaged across epochs within that condition. The primary outcome was the percentage change from baseline to post-stimulation for each channel, expressed as the relative increase or decrease versus its own baseline. Channel-level values were then summarized using the predefined anatomical/clinical groupings (SEEG: SOZ, EARLY, OTHER; scalp: ipsilateral, contralateral, midline) and aggregated by stimulated thalamic nucleus and band (slow/fast/HFA). Comprehensive power results are provided in **Supplementary Table 2**.

### Granger causality (GC) analysis

Directed interactions among bipolar SEEG channels were estimated with time-domain Granger causality on the predefined 885-ms epochs, analyzed separately for the slow and fast bands. In the standard linear framework, GC asks whether the past of signal X improves the prediction of Y beyond the past of Y alone, operationalized with bivariate vector autoregressive (VAR) models and variance-reduction F-tests.^29^

Time-domain GC and spectral GC are mathematically linked through the same VAR model: spectral GC provides a frequency-resolved decomposition, whereas time-domain GC corresponds to the frequency-integrated measure. In stimulus-locked, sub-second windows with limited samples, low-order VARs provide stable estimates; band-wise effects can then be summarized either by computing GC on band-defined epochs (as done here) or, alternatively, by averaging spectral GC within bands. We adopted the former for stability in short windows and transparency of preprocessing, consistent with standard implementations.^29^

To target frequency-specific effects in short windows, signals were zero-phase band-pass filtered into the slow and fast bands before GC estimation. This pragmatic choice is common when sample sizes per epoch are limited and low-order VARs are desired, focusing inference on physiologically relevant bands while maintaining model stability.^29,30^ Because band-pass filtering can bias GC estimates if poorly implemented, we used linear-phase, zero-lag filters applied identically to baseline and post-stimulation data and restricted interpretation to band-level summaries.

For each patient and each ordered channel pair (i,j), we fit bivariate VAR models with a maximum lag of 5 samples (≈10 ms at our sampling rate). This setting captures short-latency cortico-cortical interactions against over-parameterization in short windows, as recommended in practical GC applications.^29,30^ We extracted the F-statistic at lag 1 from the nested-model test as a scalar GC magnitude, consistent with standard routines, and computed GC matrices per epoch. Matrices were averaged across epochs to yield one baseline and one post-stimulation estimate per band; change was expressed as a percent difference relative to baseline for each directed edge and assembled into directed adjacency matrices. Node-level inflow (column-wise sum) and outflow (row-wise sum) were derived for baseline, post-stimulation, and change, and then summarized within the predefined SEEG groups (SOZ, EARLY, OTHER), as in the power spectral analysis. Cohort-level results were aggregated by stimulated thalamic nucleus and band (slow/fast). We excluded HFA from GC analysis, since convergent evidence indicates that HFA is highly local and closely tracks multiunit spiking, rather than mediating broad, long-range interactions.^31,32^ Comprehensive GC results are provided in **Supplementary Table 3**.

### Statistical analysis: power and GC flow comparisons across groups

For each metric × band × nucleus, we conducted prespecified two-group comparisons rather than omnibus tests, with two-tailed nonparametric procedures and Benjamini–Hochberg false-discovery rate (FDR) correction within each comparison family. We report FDR-adjusted *P*-values throughout, where *P* < 0.05 was considered significant. Because thalamic pairs were stimulated at 1 and 2 mA, these conditions were averaged for the primary analysis; intensity-specific results (1 mA vs 2 mA) were assessed in supplementary analyses to examine dose effects.

For SEEG power, we compared SOZ vs. EARLY, SOZ vs. OTHER, and EARLY vs. OTHER. For the primary, channel-level analysis, we used Dunn’s test across the three prespecified pairwise contrasts with FDR. As effect sizes for these contrasts, we report Cliff’s delta (δ). In the patient-level sensitivity analysis, we formed per-patient means within SOZ, Early, and Other and then applied two-tailed Mann–Whitney U for the same pairwise contrasts with FDR; effect sizes are reported as rank-biserial correlation (RBC).

For SEEG GC flows, within each band and nucleus, we tested only prespecified two-group contrasts of directed connectivity, namely:

(i) counter-direction flows between the same pair of groups (e.g., SOZ➔EARLY vs. EARLY➔SOZ);
(ii) same-source outflows to different targets (e.g., SOZ➔EARLY vs. SOZ➔OTHER);
(iii) same-target inflows from different sources (e.g., EARLY➔SOZ vs. OTHER➔SOZ).

Contrasts such as SOZ➔EARLY vs. EARLY➔OTHER were not evaluated. Edge-level values (percent change) entered two-tailed Mann–Whitney U with FDR across the set of prespecified contrasts for a given (band × nucleus). Effect sizes are reported as RBC. In the patient-level sensitivity analysis, we averaged edges per patient within each flow and applied the same Mann–Whitney/FDR framework.

Two supplementary analyses complemented the primary tests. First, motivated by thalamocortical anatomy, we compared the LIM-SOZ group vs. the non-limbic (NonLIM)-SOZ group for AN-stimulated patients. Within each band, we tested the same group (power) or the same flow (GC) across the two SOZ-location subgroups using two-tailed Mann– Whitney U with FDR across bands. Second, within each power group and each GC flow, we compared the intensities at 1 mA vs. 2 mA. When both intensities were available for the same channel (power) or the same directed pair (GC), we used the paired Wilcoxon signed-rank test with FDR. All analyses were performed using Python 3.12.4.

## Results

### SEEG power—thalamic stimulation preferentially suppresses early-propagating cortex

As a representative case (P4), **Fig. 2** displays band-limited power at baseline and after thalamic stimulation in slow, fast, and HFA bands, showing widespread post-stimulation attenuation within EARLY regions.

**Fig. 2.**
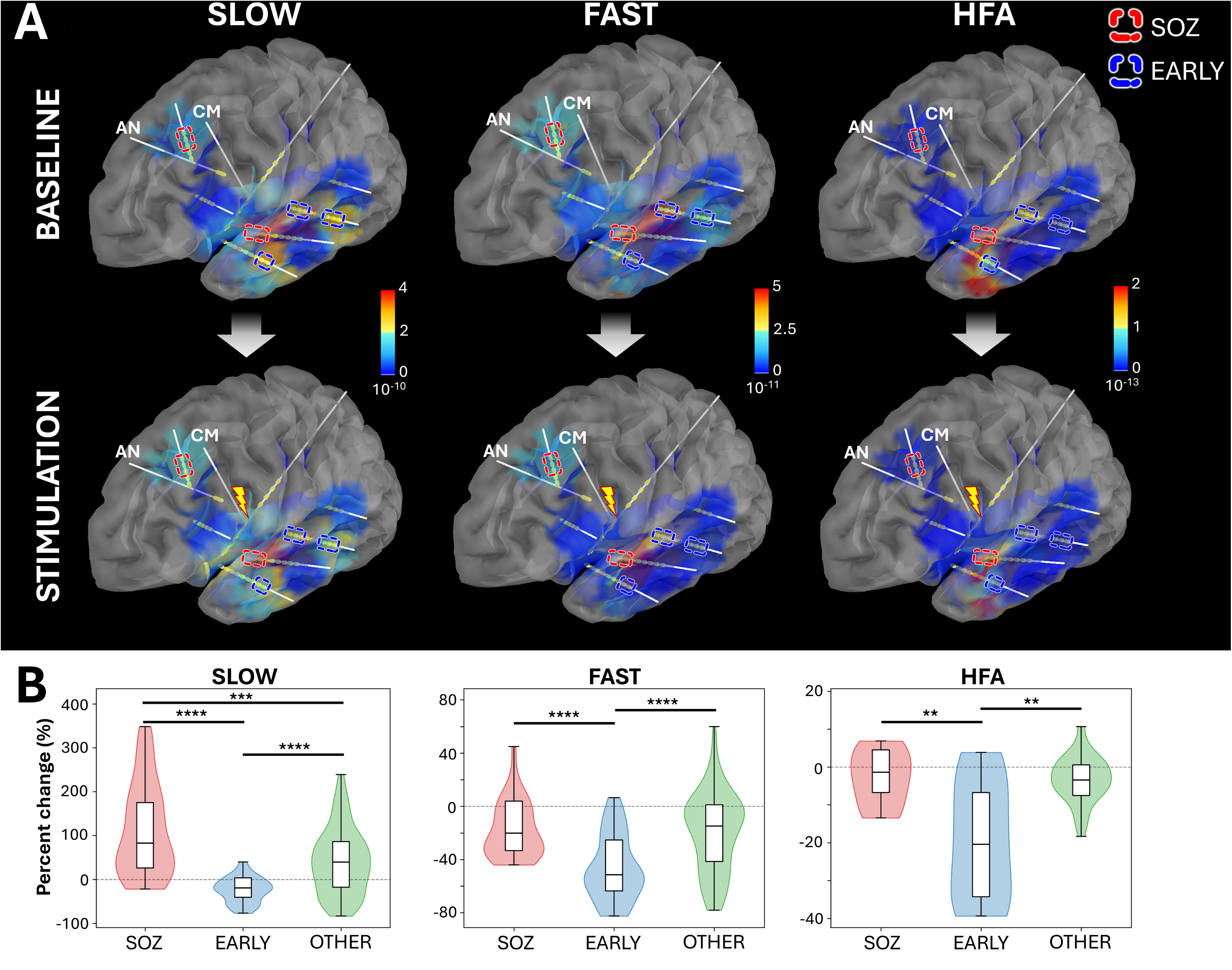
Representative case (P4): cortical power before and after thalamic CM stimulation. (A) Surface renderings of band-limited power at baseline and after single-pulse thalamic stimulation, shown for slow (3.0–12.9 Hz), fast (13.0–79.9 Hz), and high-frequency activity (80.0–299.9 Hz). Post-stimulation maps highlight focal attenuation in early-propagating cortex (EARLY) relative to SOZ and other cortical regions. (B) Distributions of percentage power change (post-stimulation relative to baseline) by cortical group (SOZ, EARLY, OTHER) for the same patient. Violin plots (with embedded box/median) summarize channel-level values; negative medians in EARLY indicate consistent reductions across bands, whereas SOZ/OTHER exhibit smaller or mixed shifts. Asterisks denote FDR-adjusted significance from two-tailed Dunn’s tests for pairwise contrasts within each band and nucleus (* *P* < 0.05, ** *P* < 0.01, *** *P* < 0.001, **** *P* < 0.0001). For visual clarity, violins are drawn after trimming outliers by the Tukey 1.5×IQR rule; all statistical tests used the full (untrimmed) data. CM = centromedian nucleus; SOZ = seizure onset zone; FDR = false discovery rate; IQR = interquartile range.

In the cohort-level analysis, EARLY was consistently and most strongly suppressed [**Fig. 3**]. For AN stimulation, EARLY decreased more than SOZ and OTHER in the slow band (*P* = 0.00054, |δ| = 0.17; *P* = 0.00047, |δ| = 0.15) and in the fast band (*P* = 2.56×10□□, |δ| = 0.22; *P* = 0.00043, |δ| = 0.15). OTHER also showed a small additional decrease relative to SOZ in the fast band (*P* = 0.049, |δ| = 0.086). In HFA, EARLY was lower than OTHER (*P* = 0.030, |δ| = 0.18). For CM stimulation, EARLY was again lower than SOZ and OTHER in slow (*P* = 5.76×10□²□, |δ| = 0.40; *P* = 6.49×10□³□, |δ| = 0.39) and fast band (*P* = 0.00011, |δ| = 0.19; *P* = 2.51×10□□, |δ| = 0.20). In HFA, both SOZ and EARLY were lower than OTHER (SOZ vs OTHER: *P* = 0.029, |δ| = 0.19; EARLY vs OTHER: *P* = 0.031, |δ| = 0.16). Together, these findings indicate that thalamic stimulation exerts the greatest acute suppression on the cortex participating in early-propagation, with effects robust across nuclei and bands and most pronounced for CM in the slow range.

**Fig. 3.**
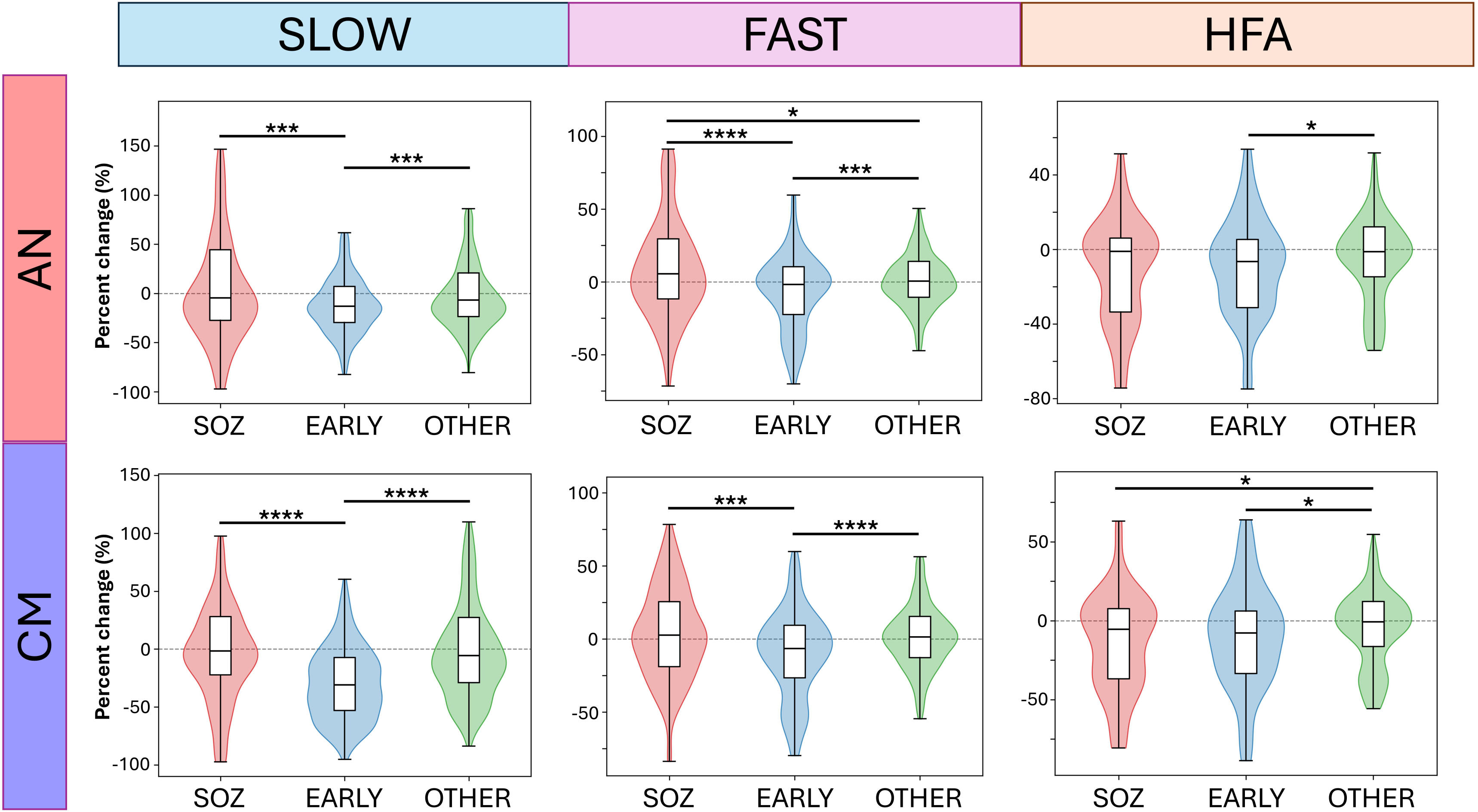
Group-level cortical power changes with thalamic stimulation. Channel-level percentage change in band-limited power (post-stimulation relative to baseline) aggregated across patients, stratified by thalamic nucleus (AN, CM), frequency band (slow: 3.0–12.9 Hz; fast: 13.0–79.9 Hz; HFA: 80.0–299.9 Hz), and cortical group (SOZ, EARLY, OTHER; ipsilateral hemisphere). Violin plots (with box/median) summarize distributions across bipolar SEEG channels; negative values indicate power attenuation. EARLY shows the most consistent attenuation in slow/fast bands for both nuclei, whereas SOZ and OTHER exhibit smaller or mixed shifts. Asterisks mark FDR-adjusted significance for pairwise contrasts within each band and nucleus using two-tailed Dunn’s tests (* *P* < 0.05, ** *P* < 0.01, *** *P* < 0.001, **** *P* < 0.0001). For visual clarity, violins are drawn after trimming outliers by the Tukey 1.5×IQR rule; all statistics used the full (untrimmed) data. AN = anterior nucleus; CM = centromedian nucleus; HFA = high-frequency activities; SOZ = seizure onset zone; FDR = false discovery rate; IQR = interquartile range.

### SEEG Granger causality—thalamic stimulation preferentially increases flow between SOZ and non-epileptic cortex

At the cohort level, directed GC changes (percent change from baseline) revealed a consistent trend toward increased inflow to the SOZ following thalamic stimulation, most prominently for NORMAL→SOZ [**Fig.4**].

**Fig. 4.**
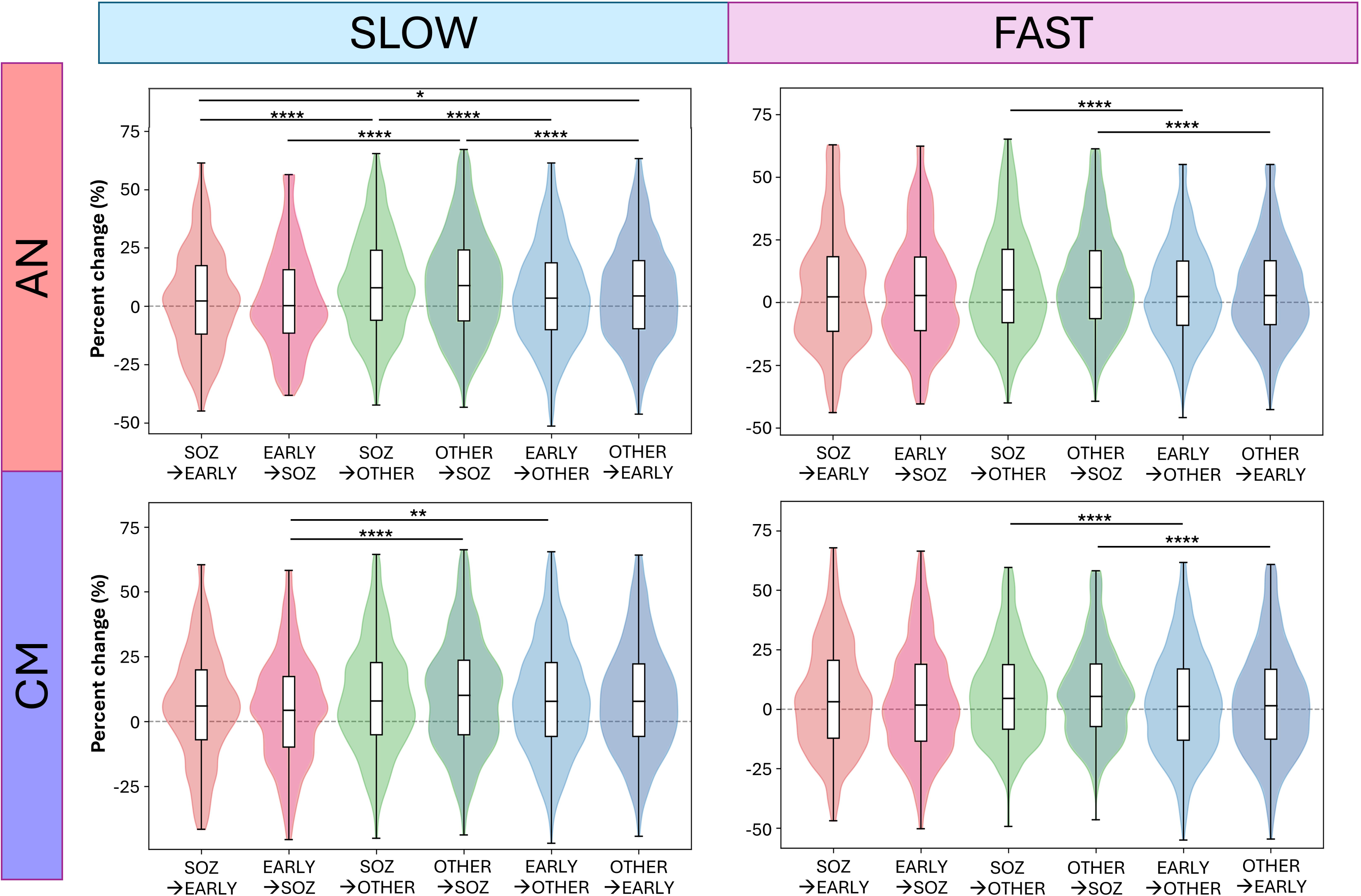
Group-level directed connectivity changes with thalamic stimulation. Percentage change in time-domain Granger causality (post-stimulation vs baseline) between bipolar SEEG channel groups, shown by thalamic nucleus (AN, CM) and band (slow: 3.0–12.9 Hz; fast: 13.0– 79.9 Hz). Rows denote directed flows (X➔Y) among SOZ, early-propagation (EARLY), and other cortex (OTHER) within the hemisphere ipsilateral to stimulation. Positive values indicate greater directed influence after stimulation. Violin plots (with box/median) summarize channel-pair (edge) distributions; notably, flows between SOZ and OTHER—particularly OTHER➔SOZ—show the most reproducible augmentation across nuclei and bands, while SOZ↔EARLY changes are smaller. Asterisks indicate FDR-adjusted significance for pairwise contrasts within each band and nucleus using two-tailed Mann– Whitney U tests (* *P* < 0.05, ** *P* < 0.01, *** *P* < 0.001, **** *P* < 0.0001). For visual clarity, violins are rendered after trimming outliers by the Tukey 1.5×IQR rule; all statistical tests used the full (untrimmed) data. SEEG = stereo-EEG; AN = anterior nucleus; CM = centromedian nucleus; SOZ = seizure onset zone; FDR = false discovery rate; IQR = interquartile range.

For AN stimulation, in slow band, OTHER➔SOZ was greater than OTHER➔EARLY (*P* = 3.36×10□□, |RBC| = 0.11) and EARLY➔SOZ (*P* = 3.55×10□□, |RBC| = 0.20). SOZ➔OTHER exceeded SOZ➔EARLY (*P* = 6.44×10□□, |RBC| = 0.17) and EARLY➔OTHER (*P* = 1.70×10□□, |RBC| = 0.12). OTHER➔EARLY was also higher than SOZ➔EARLY (*P* = 0.047, |RBC| = 0.078). For fast band, OTHER➔SOZ exceeded OTHER➔EARLY (*P* = 7.59×10□□, |RBC| = 0.11), and SOZ➔OTHER exceeded EARLY➔OTHER (*P* = 8.64×10□□, |RBC| = 0.11).

For CM stimulation, in the slow band, OTHER➔SOZ exceeded EARLY➔SOZ (*P* = 5.7×10□□, |RBC| = 0.16). In addition, EARLY➔OTHER was greater than EARLY➔SOZ (*P* = 0.0071, |RBC| = 0.11). For fast band, OTHER➔SOZ exceeded OTHER➔EARLY (*P* = 2.73×10□□, |RBC| = 0.11), and SOZ➔OTHER exceeded EARLY➔OTHER (*P* = 1.2×10□□, |RBC| = 0.10).

Taken together, these results show that thalamic stimulation amplifies GC flows both into and out of the SOZ, with the strongest and most consistent increase in OTHER➔SOZ across nuclei and bands.

### Patient-level analysis—power trends recapitulate suppression of early-propagation cortex

To verify whether channel-level patterns generalize across individuals, we repeated the analyses at the patient level. For each patient, we summarized power by taking the median percentage change within each predefined SEEG group (SOZ, EARLY, OTHER) and summarized GC by the median percentage change within each prespecified directed flow. We then applied the same within-band, nucleus-stratified pairwise tests as in the primary analysis.

For power, both AN and CM stimulation showed a tendency toward greater decreases in Early compared with SOZ and OTHER across all bands [**Supplementary Fig. 1A-B**], but none reached statistical significance given the small cohort (n=10).

For GC, patient-level flow distributions were heterogeneous and did not replicate the channel-level pattern in a consistent manner [**Supplementary Fig. 1C–D**]. These inconsistent results are expected for at least two reasons: (i) collapsing many directed edges to a single median per patient reduces sample size and can cancel opposing edges within the same flow, and (ii) between-patient heterogeneity in coverage, SOZ extent, and stimulated nucleus introduces genuine variance that is invisible at the channel/edge level.

### Anterior nucleus stimulation: limbic SOZ versus non-limbic SOZ subgroup comparison

We restricted the subgroup analysis to AN stimulation because a planned CM– frontal comparison was confounded by overlap in SOZ (six patients had frontal SOZ, of whom four also had limbic SOZ), and because CM is known to project more diffusely across associative and sensorimotor cortices than AN. In contrast, AN participates in the Papez circuit, providing a more specific limbic connectivity motif, with additional connections reaching medial/orbitofrontal regions.^19,27^ Accordingly, we contrasted patients with limbic SOZ (LIM-SOZ; n=4) against those with non-limbic SOZ (NonLIM-SOZ; n=4) under AN stimulation. Of the LIM-SOZ group, 3/4 also had frontal SOZ, and 1/4 had frontal and temporal SOZ.

For power, in the slow band, OTHER channels in the NonLIM-SOZ group showed a larger power decrease than in the LIM-SOZ group (*P* = 5.86×10□²□, |δ| = 0.43) [**Fig. 5A**]. Both groups showed a power decrease in EARLY, while there was no between-group difference. In the fast and HFA bands, Early channels in the LIM-SOZ group exhibited greater power suppression (fast: *P* = 9.61×10□□, |δ| = 0.40; HFA: *P* = 0.012, |δ| = 0.34).

**Fig. 5.**
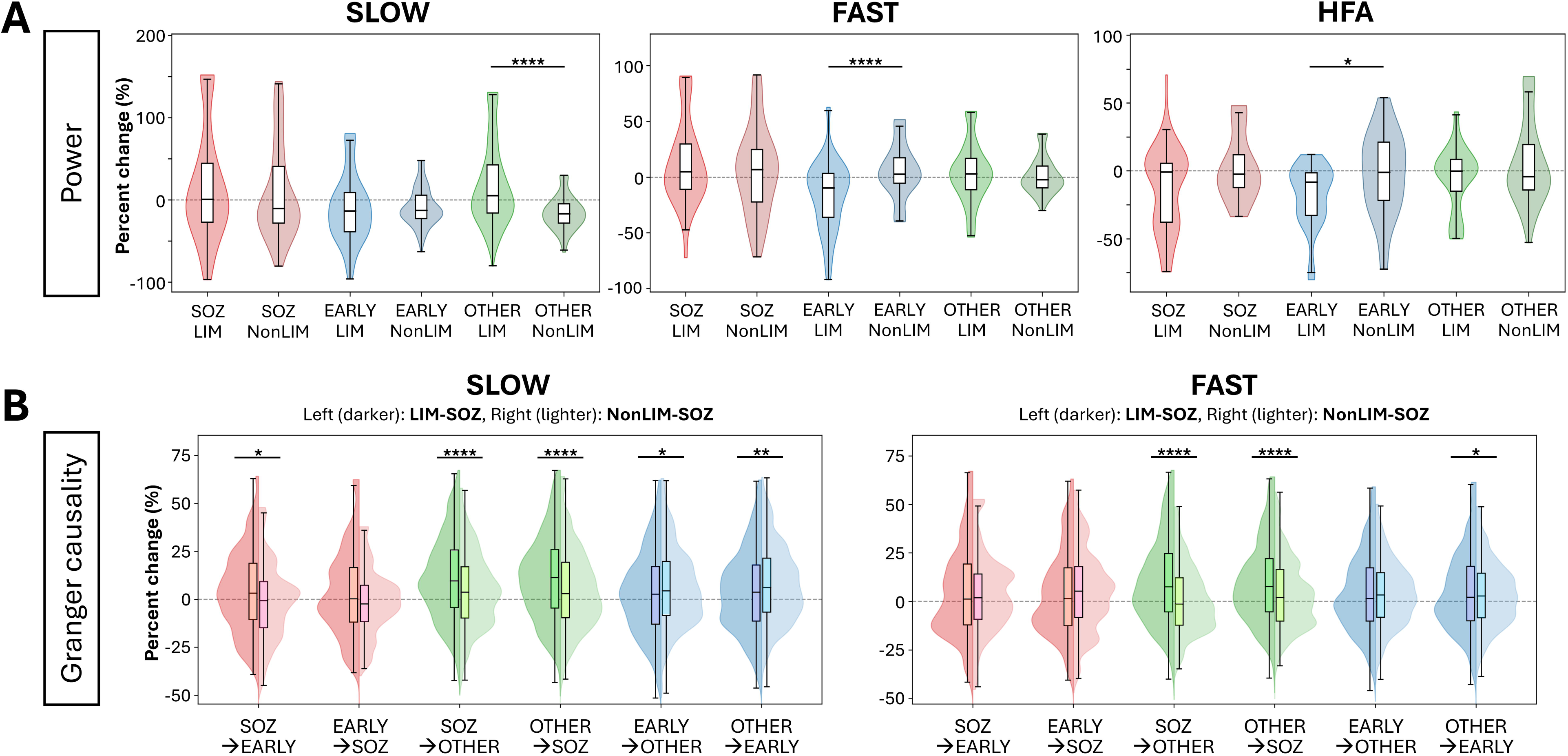
AN stimulation: comparison between limbic SOZ and non-limbic SOZ. Group-wise effects of AN stimulation (limbic: LIM vs non-limbic: Non-LIM). (A) Power change. Violin□+□box plots show percent change (post-stimulation vs baseline) in band-limited power (slow, fast, HFA) for SOZ, EARLY, and OTHER cortical groups. Left (darker) distributions correspond to LIM-SOZ group; right (lighter) to NonLIM-SOZ. In fast band and HFA, EARLY power was lower in LIM-SOZ than in NonLIM-SOZ, consistent with stronger attenuation when the limbic cortex was part of the SOZ. (B) Directed connectivity (GC) change. Violin□+□box plots show percent change in flow between cortical groups. In the slow band, LIM-SOZ demonstrated greater increases than NonLIM-SOZ for SOZ➔OTHER, OTHER➔SOZ, and SOZ➔EARLY, whereas EARLY➔OTHER and OTHER➔EARLY were higher in NonLIM-SOZ. In the fast band, LIM-SOZ again showed larger SOZ➔OTHER and OTHER➔SOZ, with OTHER➔EARLY relatively larger in NonLIM-SOZ. Asterisks indicate FDR-adjusted significance for pairwise contrasts within each band and nucleus using two-tailed Mann–Whitney U tests (* *P* < 0.05, ** *P* < 0.01, *** *P* < 0.001, **** *P* < 0.0001). For visual clarity, violins are drawn after trimming outliers by the Tukey 1.5×IQR rule; all statistical tests used the full (untrimmed) data. AN = anterior nucleus; SOZ = seizure onset zone; HFA = high-frequency activities; FDR = false discovery rate; IQR = interquartile range.

For GC, in the slow band, flows into and out of SOZ were higher in LIM-SOZ than NonLIM-SOZ for OTHER➔SOZ (*P* = 1.6×10□□, |RBC| = 0.18), SOZ➔OTHER (*P* = 3.8×10□□, |RBC| = 0.16), and SOZ➔EARLY (*P* = 0.038, |RBC| = 0.15) [**Fig. 5B**]. By contrast, OTHER➔EARLY and EARLY➔OTHER were higher in NonLIM-SOZ (*P* = 0.0041, |RBC| = 0.10; *P* = 0.021, |RBC| = 0.080). In the fast band, OTHER➔SOZ and SOZ➔OTHER were higher in LIM-SOZ (*P* = 4.30×10□□, |RBC| = 0.24; *P* = 1.58×10□¹³, |RBC| = 0.32), whereas OTHER➔EARLY was higher in NonLIM-SOZ (*p* = 0.038, |RBC| = 0.076).

Taken together, AN stimulation shows a limbic-selective modulation: when the SOZ includes limbic cortex, Early cortex power is preferentially suppressed, particularly at higher frequencies, and bidirectional SOZ↔OTHER GC increases, whereas non-limbic SOZ exhibits stronger OTHER↔EARLY cortico-cortical reweighting.

### Stimulation intensity comparison—GC flows scale with intensity, while power effects remain inconsistent

We evaluated whether thalamic stimulation intensity altered the magnitude of network responses beyond the primary effects reported above. For power, intensity effects were small and not consistent across bands. With AN stimulation, only HFA showed a difference, with greater decreases at 1 mA than 2 mA (*P* = 0.0015, |RBC| = 0.18) [**Supplementary Fig. 2A**]. With CM stimulation, slow-band decreases were slightly larger at 1 mA in SOZ and EARLY (*P* = 0.028, |RBC| = 0.14; *P* = 0.0020, |RBC| = 0.14), whereas in HFA the OTHER group showed larger decreases at 2 mA (*P* = 0.0029, |RBC| = 0.16) [**Supplementary Fig. 2B**].

In contrast, GC showed a clear intensity scaling. With AN stimulation, 2 mA produced higher SOZ↔OTHER flows in both bands (slow: SOZ➔OTHER *P* = 5.21×10□□, |RBC| = 0.18; OTHER➔SOZ *P* = 3.61×10□□, |RBC| = 0.19; fast: SOZ➔OTHER *P* = 4.36×10□□, |RBC| = 0.20; OTHER➔SOZ *P* = 1.06×10□□, |RBC| = 0.17) [**Supplementary Fig. 2C**]. With CM stimulation, all six flows were higher at 2 mA in both slow (SOZ➔EARLY, EARLY➔SOZ, SOZ➔OTHER, OTHER➔SOZ, EARLY➔OTHER, OTHER➔EARLY: *P* = 3.94×10□¹¹, 1×10□¹³, 1.88×10□□, 4.3×10□¹², 3.65×10□¹□, 6.88×10□□; |RBC| = 0.45, 0.52, 0.20, 0.27, 0.19, 0.17, respectively) and fast bands (*P* = 5.14×10□¹¹, 2.36×10□¹¹, 5.14×10□¹¹, 2.36×10□¹¹, 2.64×10□□, 2.05×10□¹¹; |RBC| = 0.44, 0.46, 0.26, 0.27, 0.16, 0.20, respectively) [**Supplementary Fig 2D**]. Thus, while power differences were modest and band-limited, GC showed robust, direction-specific increases at higher intensity, especially under CM stimulation.

Importantly, these intensity effects do not alter the direction of our main findings, but rather scale their magnitude. This supports our choice to average intensities in the primary analyses, while acknowledging that 2 mA tends to enhance corticocortical GC flows.

### Scalp EEG power

In the eight patients with concurrent scalp EEG, we assessed band-limited power changes during thalamic stimulation. With AN stimulation, in the fast band, there was a trend toward greater decreases ipsilateral to stimulation than at the midline, which did not survive multiple-comparisons correction (*P* = 0.051, |δ| = 0.27) [**Supplementary Fig. 3A**]. With CM stimulation, no between-group differences were detected across all bands [**Supplementary Fig. 3B**]. Across nuclei, hemispheres, and frequency bands, scalp power tended to show a diffuse decrease during stimulation, without a reliable lateralized effect.

## Discussion

In a pediatric–adolescent SEEG cohort, we demonstrate that repetitive SPES of the anterior or centromedian thalamic nuclei modulates cortical network dynamics within 1 s. First, band-limited power in early-propagating cortex decreases, almost consistently across slow/fast/HFA bands. Second, directed connectivity is enhanced modestly at the network level, with the most reproducible augmentation between SOZ and non-SOZ cortex, especially for influence toward SOZ. These patterns held despite heterogeneous implantation plans and seizure semiology, and they were most evident when AN was stimulated in patients with limbic SOZ. Taken together, these results support a network-level mechanism whereby thalamic input perturbs cortico-cortical interactions in a way that attenuates propagation from SOZ to early-propagating regions. Propagation could also be limited if stimulation engages the thalamic reticular nucleus (TRN), leading to inhibition of thalamic relays and reduced thalamocortical drive.^33^ This interpretation is consistent with classic centrencephalic theories of seizure spread as well as with modern circuit models.^34,35^

### Interictal epileptogenic networks and therapeutic neuromodulation

Interictal network organization in focal epilepsy reflects a dynamic interplay between seizure-capable and surrounding cortex rather than a single static focus. The interictal suppression hypothesis posits that the non-epileptic cortex actively suppresses epileptogenic regions.^35^ In parallel, the neural fragility framework formalizes how small perturbations can destabilize epileptogenic networks, offering a systems-level lens on seizure propensity.^36^ For patients in whom resection of the epileptogenic zone is not feasible, evidence is accumulating that thalamic stimulation can modulate epileptogenic networks, with emerging clinical benefit.^4–12,37^

Within this system’s framing, nucleus-specific anatomy provides a mechanistic context. AN is embedded in the Papez/limbic circuit and projects to hippocampus, cingulate, and orbitofrontal networks, whereas CM participates in fronto-parietal and motor association loops and exerts diffuse modulatory influence across widespread cortex.^38,39^ In our cohort, despite heterogeneous SOZ topographies, thalamic stimulation produced convergent signatures (EARLY power decrease; increased non-SOZ↔SOZ flow), consistent with broad network-level modulation by both nuclei. Notably, with AN stimulation in limbic SOZ, effects were accentuated, likely reflecting closer alignment between the stimulation target and limbic connectivity; all limbic-SOZ cases also had frontal involvement, further matching known AN projection to limbic–fronto-cingulate cortices.^18,27,40^

### Power and directed flow changes in epileptogenic networks

Band-limited power tracks local population activity and network engagement. Intraoperative human data show that AN stimulation can reduce beta/low-gamma power while increasing large-scale coupling on scalp EEG.^19^ Our finding that EARLY power declines after thalamic stimulation fits with reduced recruitment of early-propagating regions during the interictal window. Although scalp EEG power suggested broad reductions at all frequency bands, the sample size limited statistical support.

For directed connectivity, GC flow increased modestly overall, with the clearest effects along the SOZ↔Other axis, especially for influence toward SOZ. Enhanced directed coupling after thalamic stimulation can coincide with network-stabilizing control at the circuit level, given TRN-mediated gating and extrathalamic inhibitory inputs that regulate thalamocortical throughput and cortical synchronization.^33,39,41^ This framework also aligns with centrencephalic notions in which thalamocortical relays act as propagation hubs that can be disrupted by recruitment of inhibitory gates rather than by direct suppression at a cortical source.^33,42^ In keeping with thalamic microcircuit anatomy, predominant thalamocortical drive coupled with TRN-mediated gating offers a straightforward explanation of how stimulation could either facilitate or prevent propagation depending on the balance of recruitment. In human and animal data, increases in network coordination metrics (including coherence and GC) can reflect effective stabilization rather than excitation per se, depending on where in the loop inhibition acts.^19,43,44^ GC indicates directed relationships, while it does not specify whether effects are excitatory or inhibitory.^45^ We therefore interpret the non-SOZ➔SOZ connectivity increase as consistent with a stabilizing influence, potentially cortico-cortical control via the thalamus.

Frequency specificity offers additional clues. Slow/fast rhythms support large-scale cortico-cortical coordination and were most consistently modulated by thalamic stimulation, whereas HFA—sensitive to local excitability—can be focal and variable across patients.^46–48^ It is therefore plausible that sub-second thalamic drive preferentially reshapes global slow/fast coordination, while HFA exhibits heterogeneous responses.^19,38^

### Emergence of thalamic stimulation evidence and translational implications

Evidence increasingly points to the thalamus as a practical target for modulating epileptic networks. Prior human study showed that the thalamic nuclei engaged by cortical SOZ during stimulation were also those with ictal involvement.^18^ Our results are consistent with intraoperative human stimulation studies showing that AN stimulation reduces 20–70 Hz power and increases diffuse connectivity on the scalp EEG.^19^ Longitudinal neuromodulation studies further suggest that effective thalamic DBS/RNS can reduce interictal spikes and reshape network organization over time.^28,37^

Our SEEG-based paradigm adds temporal specificity: by resolving sub-second dynamics, we capture the immediate network response to thalamic stimulation—the timescale on which DBS/RNS pulses act. The observed results—reduced EARLY power with the most consistent increase in non-SOZ➔SOZ directed influence—likely reflect processes repeatedly recruited during therapeutic stimulation. As a working model, we propose that thalamic stimulation recruits inhibitory inputs from non-SOZ cortex onto the SOZ, thereby reducing SOZ drive to early-propagating regions; the resulting weaker SOZ → EARLY propagation accounts for the observed power decreases in EARLY [**Fig. 6**].

**Fig. 6.**
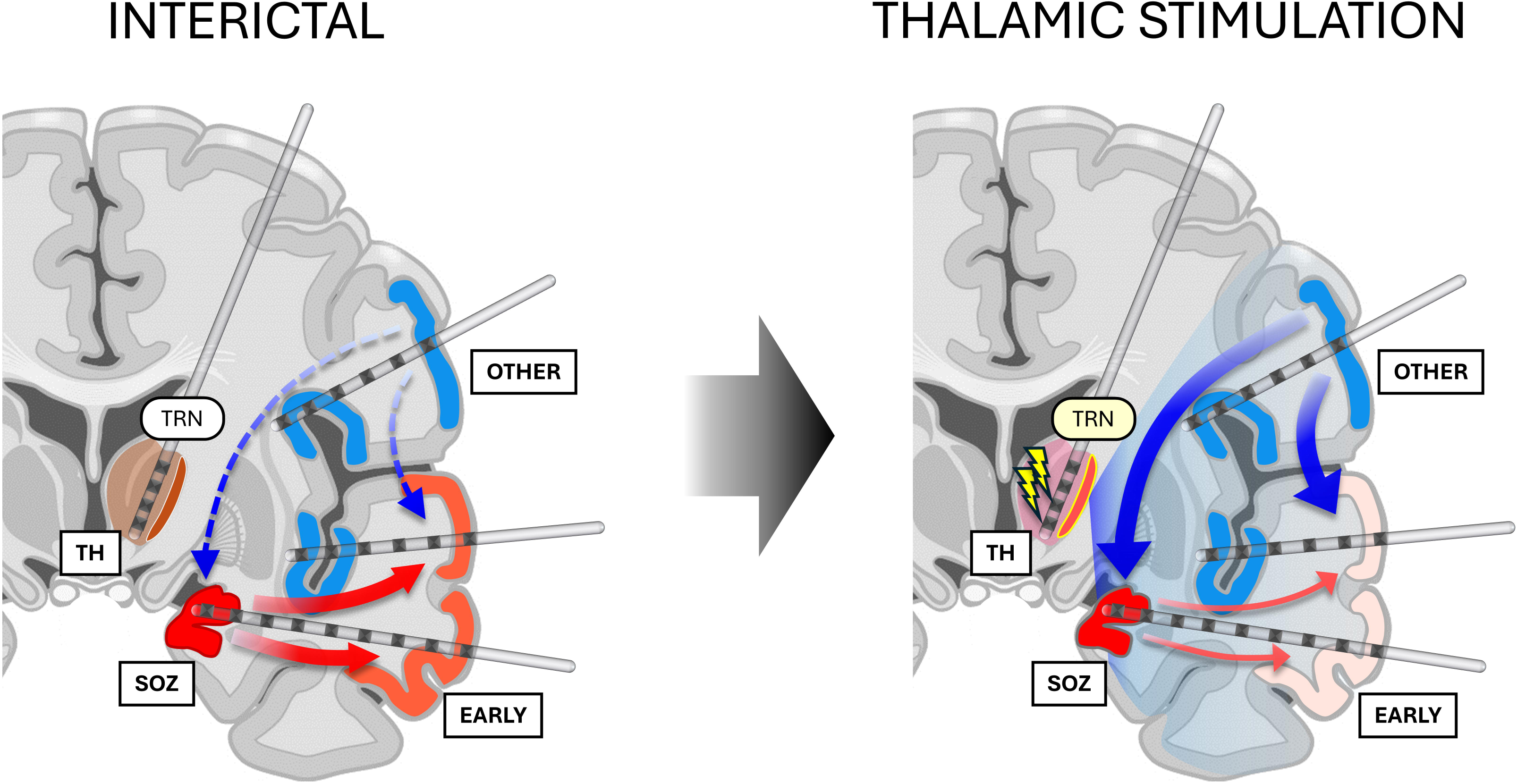
Proposed schematic model of network changes with thalamic stimulation. Conceptual summary of the proposed mechanism underlying thalamic stimulation effects on the epileptogenic network during the interictal state. Left, interictal: activity in SOZ tends to propagate to early-propagating cortex (EARLY), contributing to elevated power in EARLY. Right, during thalamic stimulation: brief thalamic (TH) input possibly engages the thalamic reticular nucleus (TRN), which gates thalamic relays and recruits broader cortico-cortical control, yielding an effectively suppressive influence from non-SOZ cortex toward SOZ and thereby reducing effective propagation from SOZ to EARLY. In this framework, EARLY power decreases because EARLY is less recruited by the SOZ drive. Arrows illustrate hypothesized direction and relative strength of interactions among SOZ, EARLY, and OTHER SOZ = seizure onset zone.

This suggests two translational paths. First, network-guided neuromodulation could serve as intra- or peri-implant acute physiological biomarkers: preferential enhancement of non-SOZ➔SOZ influence and concomitant attenuation of EARLY power could aid target selection and parameter titration. Second, these markers are testable in closed-loop systems: if this short-term signature reliably predicts a long-term improvement, devices could adaptively adjust toward settings that maximize it, similar to network reorganization associated with clinical response in cortical RNS.^49^ In short, by linking thalamic stimulation to rapid, measurable shifts in cortico-cortical dynamics, our results support network-guided strategies and define clear, physiology-based endpoints for prospective trials that integrate anatomy, acute SEEG responses, and clinical outcomes.^37^

### Methodological Considerations and Future Directions

These results should be interpreted with caution. First, this study is constrained by a small, clinically heterogeneous cohort. Although epilepsy onset occurred in childhood, the ages at SEEG evaluation varied substantially, which could influence network properties and their responsiveness to thalamic stimulation. This limits generalizability and increases between-patient variance in network organization.

Second, spatial sampling was dictated by clinical hypotheses, so parts of the epileptogenic network may have been unsampled. The anatomical distribution of channels classified as EARLY or OTHER was based solely on EEG evaluation and was not fully stratified by lobe or by their functional distance and orientation relative to the stimulated nucleus. Moreover, SOZ labels were assigned by expert review and varied across patients, introducing potential misclassification and sampling bias. In future work, implementing connectivity-informed distances between the stimulated thalamic nucleus and each cortical contact—using subject-specific MRI tractography to estimate thalamocortical pathways— could relate effects to connection strength and tract pathways, thereby strengthening interpretability and reducing sampling bias.

Finally, we did not correlate stimulus-locked network changes with long-term outcomes under DBS or RNS, leaving the link from acute modulation to durable clinical benefit inferential. In addition, cortical SPES preceded thalamic SPES in our workflow and may have induced short-term plasticity that we could not quantify. Prospective studies with larger, anatomically stratified sampling across AN and CM targets, systematic parameter mapping (intensity, frequency, pulse width), and integration with longitudinal outcomes will be essential to validate predictive value.

## Acknowledgements

The authors are deeply grateful to the patients who participated in this study and gave consent for the collection of their data for the advancement of knowledge and for a better future for epilepsy patients. We are indebted to Joyce H. Matsumoto, Lekha M. Rao, Shaun A. Hussain, Rajsekar R. Rajaraman, Samuel Ahn, Maria Garcia Roca, Richard Le, Patrick Wilson, Cesar Dominguez, and Jimmy C Nguyen for their assistance with the study and sample acquisition.

## Supplementary material

Supplementary material is available at Brain online.

## Data availability

The data and analysis code that support the findings of this study will be available upon publication. The EEG data are not publicly available due to privacy or ethical restrictions.

---

**Supplementary Table 1.** Comprehensive SEEG electrode information

**Supplementary Table 2.** Power spectral analysis results

**Supplementary Table 3.** Granger causality analysis results

**Supplementary Fig. 1.** Patient-level results

**Supplementary Fig. 2.** Stimulation intensity comparison

**Supplementary Fig. 3.** Scalp power spectral results

