## Supplementary figures and images for "Short-Term Modulation of Epileptic Network with Low-Frequency Thalamic Stimulation"

### Suppl-Fig1

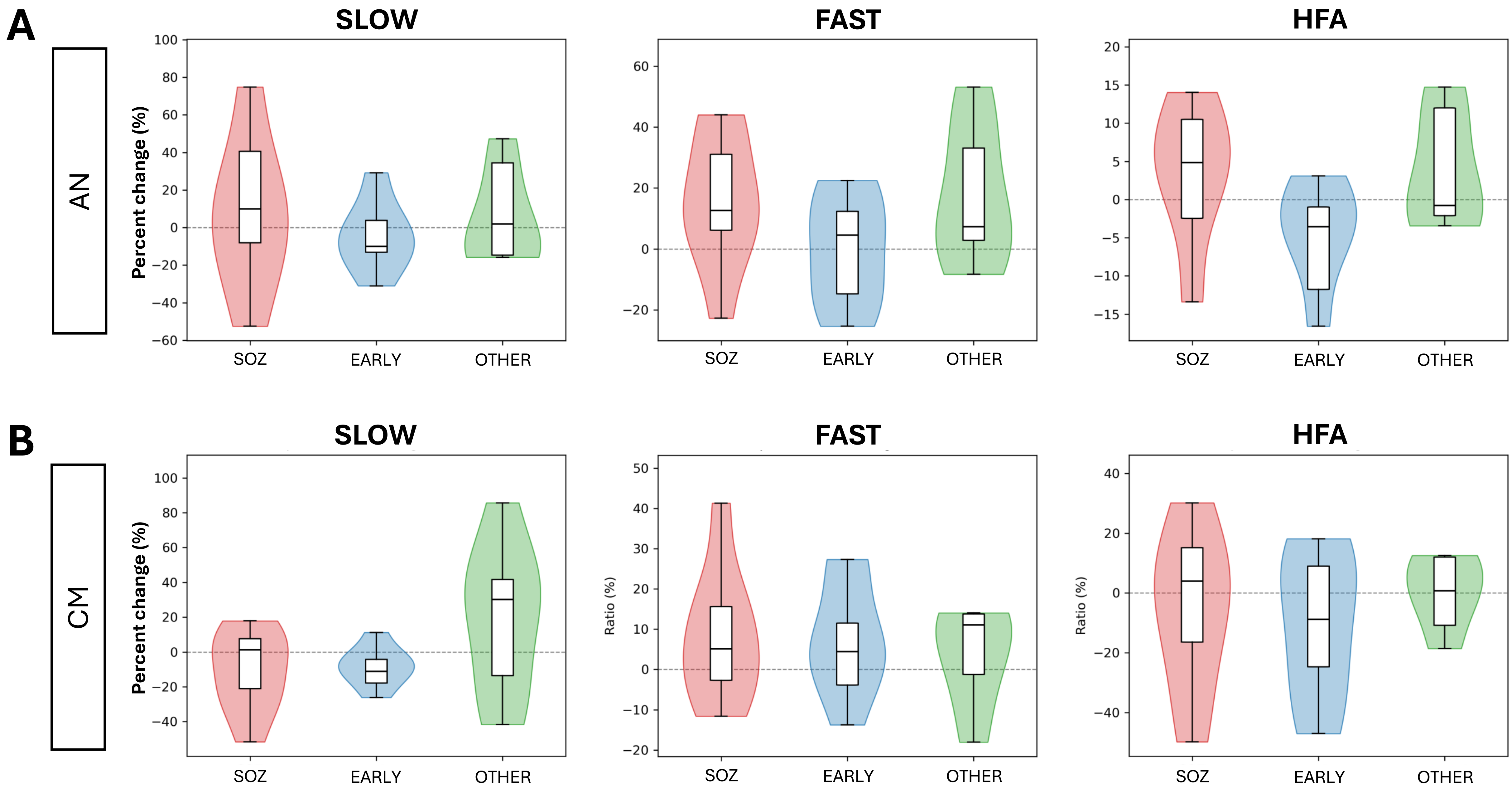

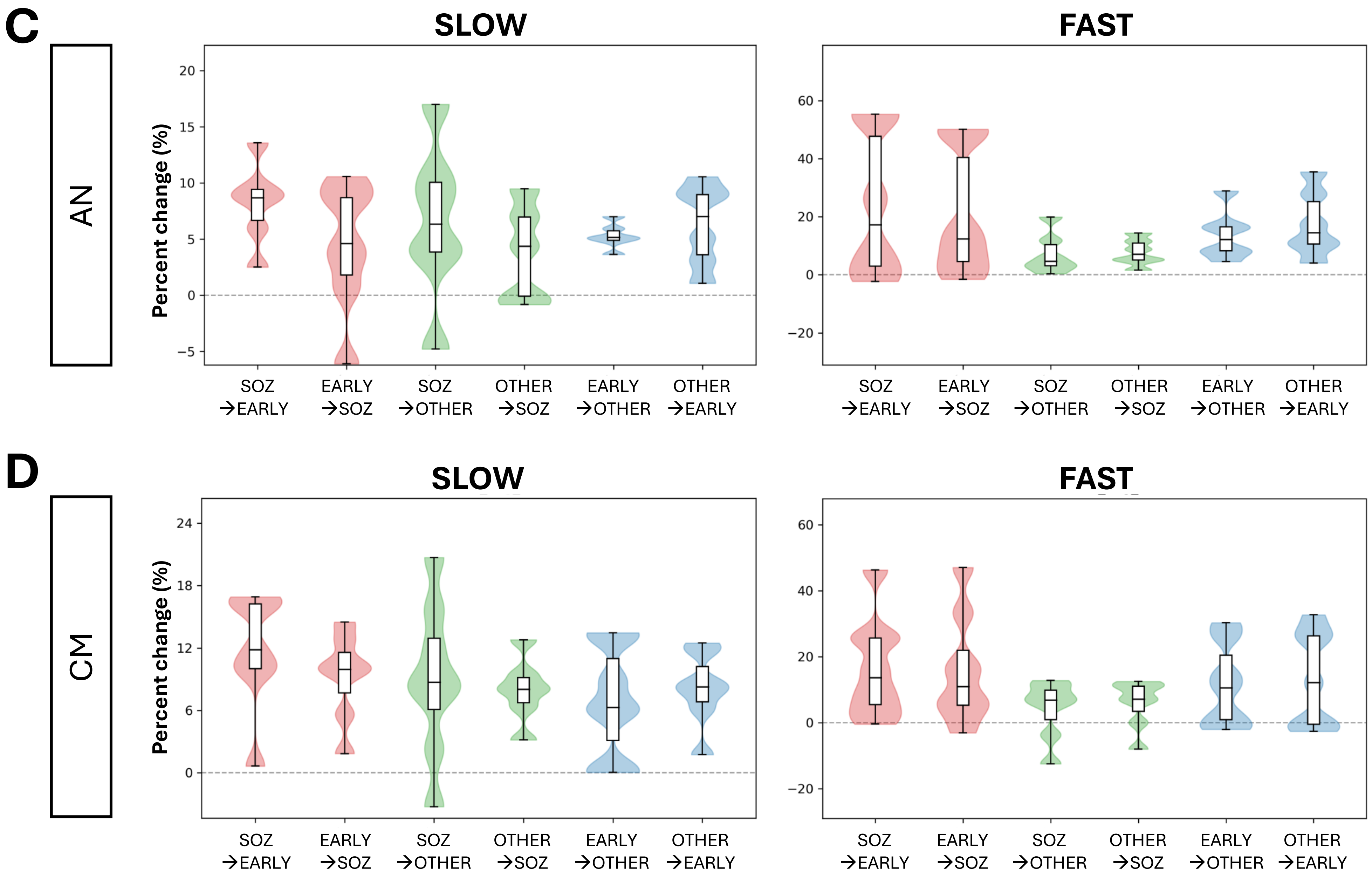

### Suppl-Fig2

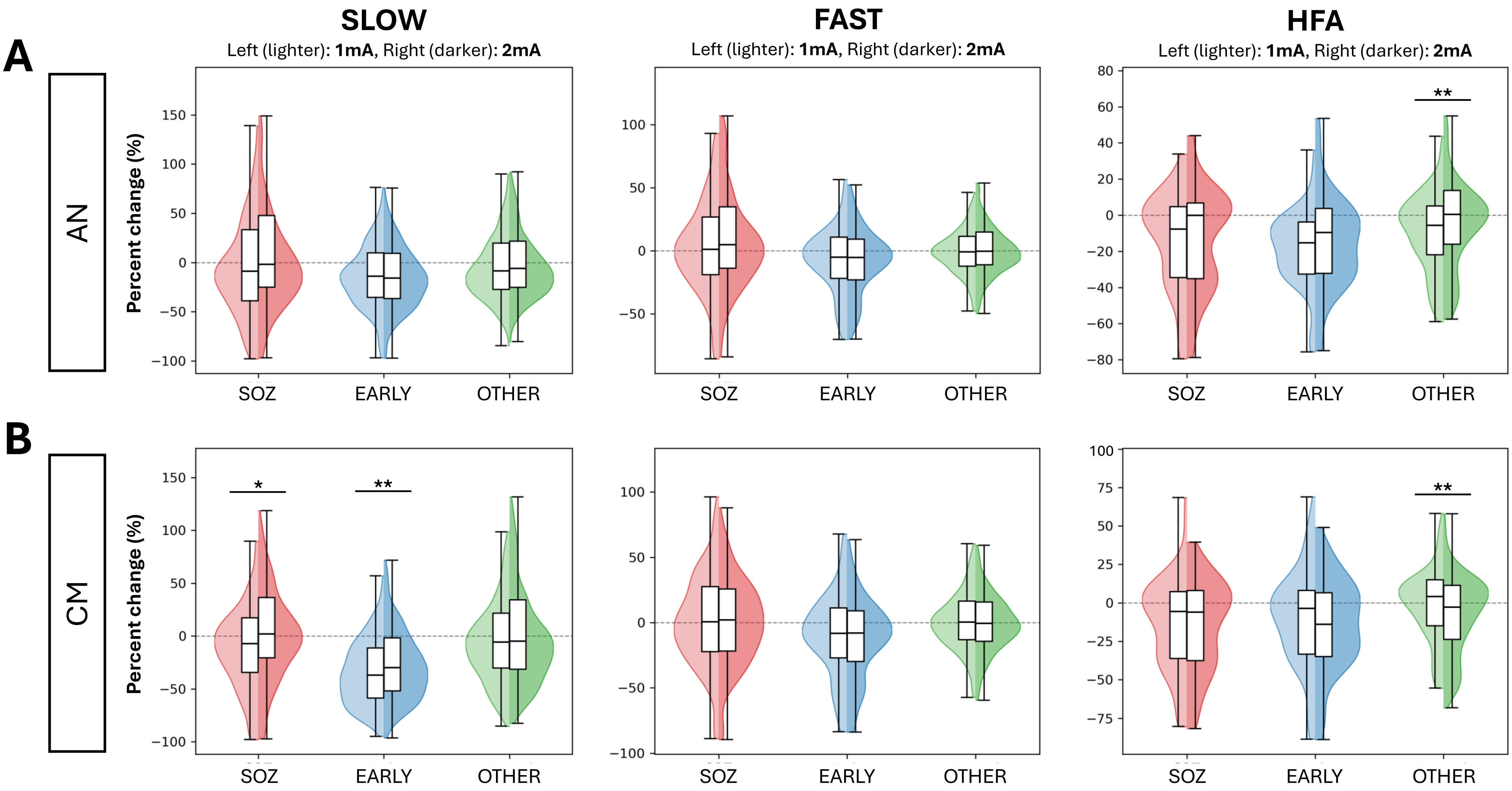

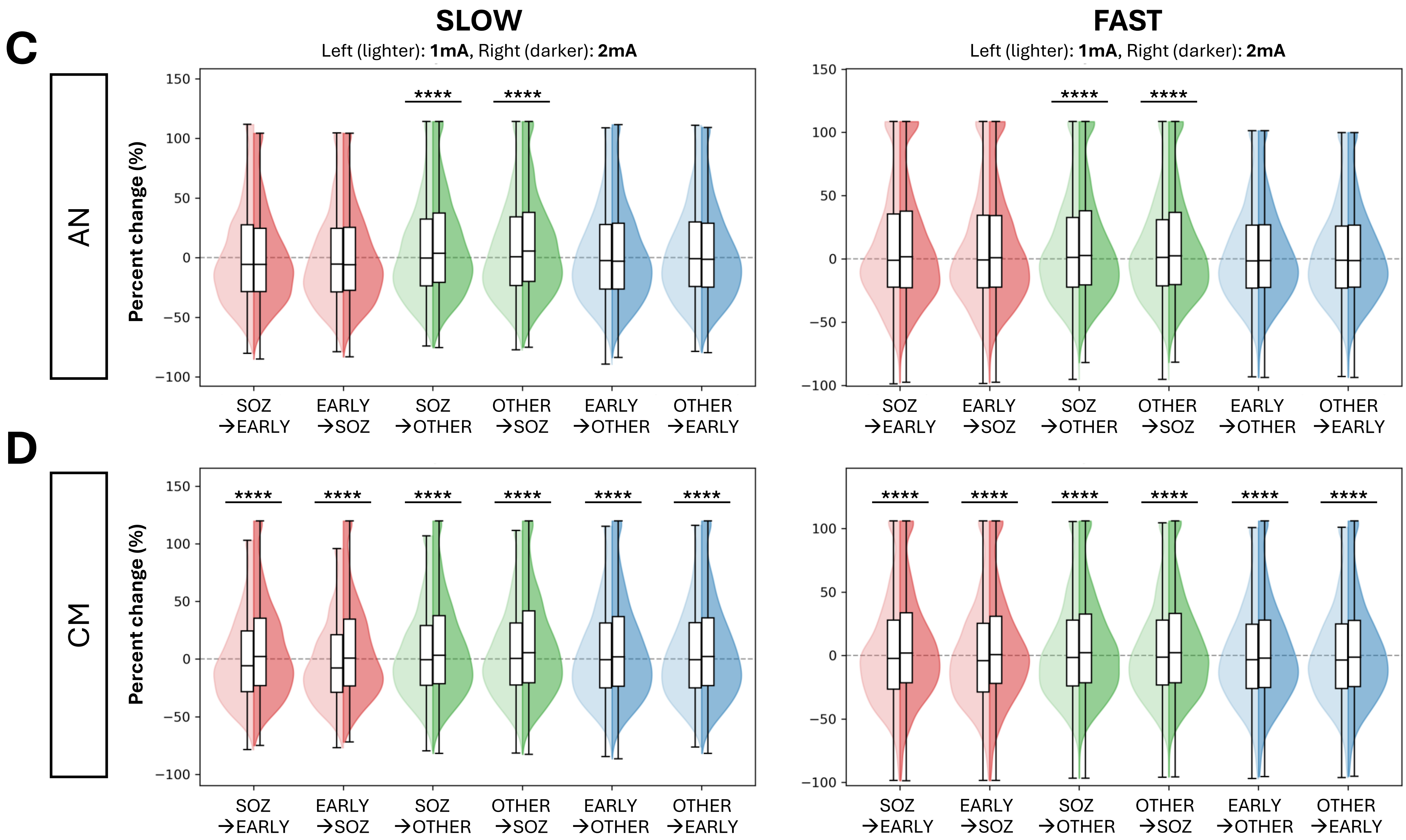

### Suppl-Fig3

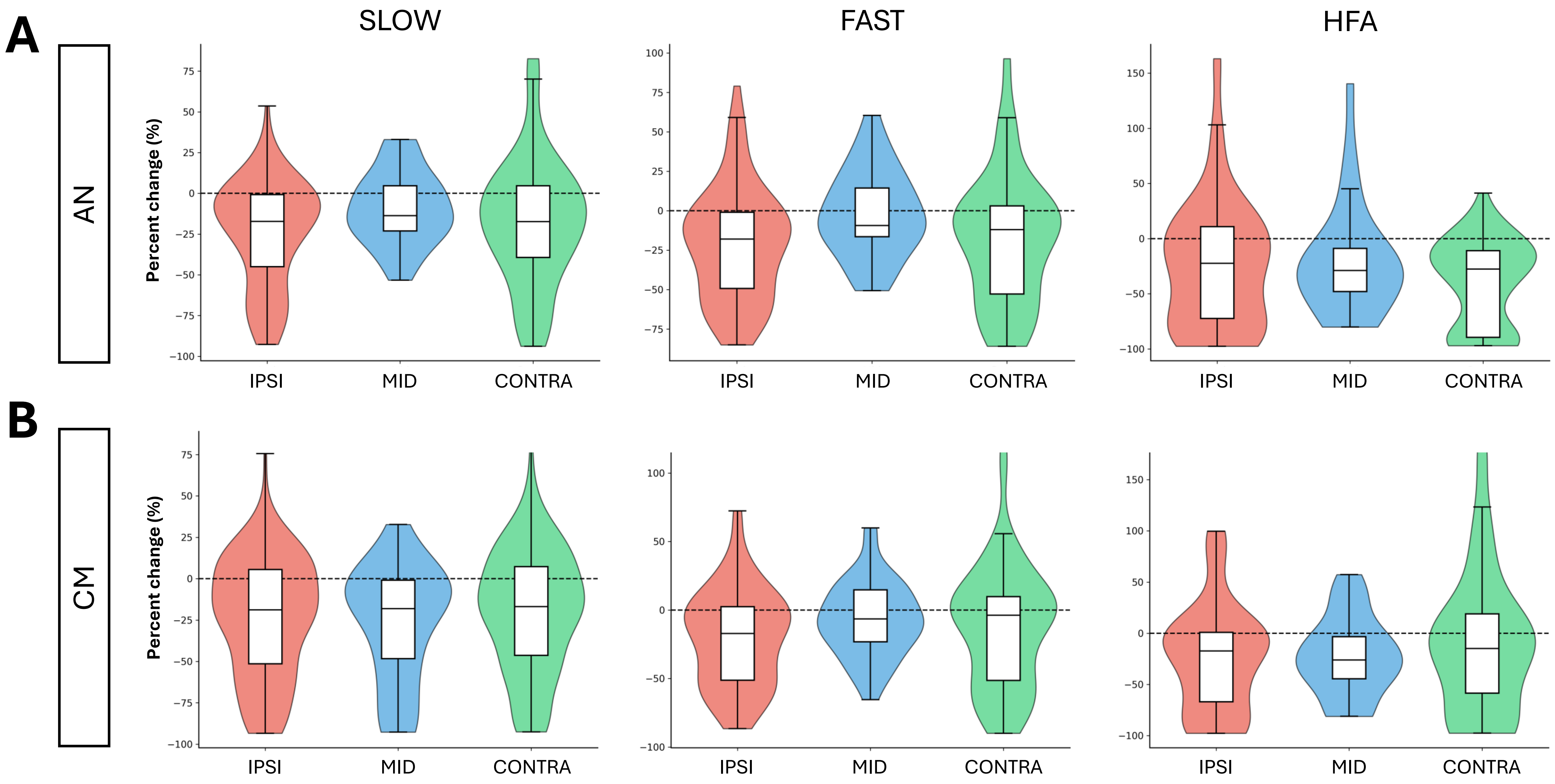
